# Elevated Cardiovascular Biomarkers Following Hypertensive Disorder of Pregnancy

**DOI:** 10.1101/2024.08.06.24311582

**Authors:** Austin M. Gabel, Lindsay Cheu, Mindy Pike, Kelsey L Olerich, Alisa Kachikis, Stephen A McCartney, Raj Shree

## Abstract

Hypertensive disorder of pregnancy (HDP) is associated with an increased risk for later-life cardiovascular disease (CVD). Whether the HDP pregnancy itself confers risk towards CVD later in life is suggested in several epidemiologic studies. Given this connection and that the HDP exposure itself may play a role, understanding whether markers associated with cardiovascular risk vary based on HDP history in the years following pregnancy may assist with risk stratification and development of targeted interventions. We measured 77 proteins (CVD-associated and inflammatory markers) in n=22 individuals with a history of HDP and n=43 matched controls with no HDP history at a median of 4 years after pregnancy. Several CVD-associated proteins (fibrinogen, fetuin-A, L-selectin, and alpha-1-acid glycoprotein) were significantly elevated, by orders of magnitude, in individuals with a history of HDP compared to normotensive pregnancies (all p<0.0001). In multivariable linear regression models controlling for age, body mass index, chronic hypertension, and diabetes, a history of HDP remained associated with higher levels of CVD-associated proteins (all p<0.0001). We clustered samples based on global patterns of CVD protein expression and found a significant difference in CVD protein expression patterns between post-Normal and post-HDP samples. Conversely, differences in circulating inflammatory markers were largely insignificant or more subtle than that observed with the CVD-associated proteins. Identification of biomarkers associated with CVD in the intervening years after HDP but before evident CVD is critical to understanding post-HDP cardiovascular risk to provide insight for the development of therapeutic interventions that mitigate CVD event risk in this high-risk population.

## INTRODUCTION

Cardiovascular disease (CVD) is the leading global cause of death^1^ and women experience significantly higher morbidity and mortality from CVD compared to men.^2^ Reproductive factors may partially explain this sexual dimorphism.^2,3^ Several epidemiological studies demonstrate a strong link between hypertensive disorder of pregnancy (HDP), such as preeclampsia, and an increased risk for later-life CVD, including coronary artery disease, myocardial infarction, heart failure, and stroke.^4–9^ This risk is further multiplied by recurrent episodes of HDP and preterm HDP. The mechanisms contributing to increased CVD risk following HDP are poorly understood. Although underlying risk factors (e.g., obesity, diabetes, chronic hypertension), likely play a strong role,^10^ epidemiologic and some translational data suggests that the HDP pregnancy itself may be an independent risk factor for CVD development.^5,7,8,11^ Although national societies include history of HDP in their algorithm for CVD risk classification,^12^ formal evidence-based guidelines for long-term screening, monitoring, or interventions do not exist due to a poor understanding of the clinical and molecular trajectory following HDP but before evident CVD.^13^

Although the clinical symptoms of HDP overwhelmingly resolve following delivery, studies demonstrate that the characteristic vascular dysfunction of HDP persists, over the short (1-3 months) and long-term period (up to 15 years after delivery).^14–17^ Individuals with a history of HDP have attenuated cerebral vascular reactivity,^18^ increased arterial stiffness,^19–21^ and increased carotid-intima thickness,^22–24^ an early measure of atherosclerosis, years after delivery. Additionally, subclinical cardiac impairment can be detected after HDP.^25,26^ A key study of a cohort just 2 to 7 years (median 3 years) after their first pregnancy noted a higher risk of hypertension in those with recent pregnancies complicated by HDP, with over a 4-fold greater risk when HDP was accompanied by indicated preterm birth.^27^ Another study found that elevated third trimester blood pressure was associated with dyslipidemia 10-14 years after pregnancy.^28^ Although clinical CVD largely occurs many years after HDP, subclinical changes are likely evident prior to this. Thus, the intervening years present a unique time for risk-stratification and consideration of therapeutics.

Mechanistic studies have recently begun to understand the pathways contributing to persistent vascular changes following HDP that may ultimately lead to CVD.^29,30^ As inflammation is an overlapping phenomenon between HDP and CVD,^31–36^ the role of inflammatory markers in explaining this connection is also of interest.^37^ Additional pathways that may contribute to altered vascular function, however, remain understudied. Critical to our understanding of long-term CVD risk following HDP are investigations into molecular signatures in the years after the HDP pregnancy. We sought to assess levels of circulating plasma proteins associated with inflammation and cardiovascular risk among those with a prior history of HDP compared to those with no HDP history. We hypothesize that CVD-associated and inflammatory proteins will be elevated in those with HDP history compared to those with no HDP history.

## DATA AVAILABILITY

De-identified clinical meta data and raw values for protein concentrations have been made publicly available at Dryad and can be accessed at https://doi.org/10.5061/dryad.rn8pk0pmc.

## METHODS

### Study Participants

Participants were prospectively recruited and underwent written informed consent through an approved Institutional Review Board protocol (STUDY00001636). Among non-pregnant participants that we recruited, we identified n=22 samples from participants with a history of HDP in a prior pregnancy (cases). Controls (n=43) were identified by matching cases (approximately 1:2) to interval since most recent pregnancy. We did not exclude participants if they had chronic hypertension or diabetes (pre-existing or history of gestational diabetes) from either group. Participants were not pregnant and presented for outpatient peripheral blood draws and were free of any acute illnesses at the time of sampling. AG, MP, and RS had full access to all the data and take responsibility for its integrity and the data analysis.

### Sample Collection and Processing

Peripheral blood was collected in acid citrate dextrose solution A-vacutainer tubes and underwent processing (unfrozen) within 24 hours (majority within 12 hours). Plasma was isolated by centrifugation of whole blood for 10 minutes at 1500 g and stored at −80°C. All samples were submitted for multiplex assay together and were run in the same batch.

### Multiplex Assay

We collected single time point plasma samples from each participant and submitted aliquots for multiplex LASER assay protein quantification.^38^ This multiplex technology relies on color-coded polystyrene beads. This minimizes sample volume needs while maximizing the number of measures per sample. Bead coloration is achieved by utilizing different concentrations of red and infrared fluorophore dyes to create 100 uniquely colored bead sets which can be combined within the same assay well. Each analyte is distinguished from the other because they are bound to differently colored/fluorescent beads. The bead analyzer (Bio-Plex 200) includes a dual-laser system and a flow-cytometry system. One laser activates the fluorescent dye within the beads which identifies the specific analyte. The second laser excites the fluorescent conjugate (streptavidin-phycoerythrin) that has been bound to the beads during the assay. The amount of the conjugate detected by the analyzer is in direct proportion to the amount of the target analyte. The results are quantified according to a standard curve. Individual protein measures with values out of range based on assay parameters were excluded.

In all participants, we measured a total of 77 proteins encompassing three distinct custom protein panels – CVD associated proteins (n=15), immune-related cytokines (n=47), and soluble cytokine receptors (n=15) (**Table 1**).

**Table 1:** Circulating proteins measured in plasma via multiplex assay.

| Protein Panel | Individual Proteins Measured |
| --- | --- |
| *CVD-associated Proteins* | BNP, NTproBNP, CK-MB, CXCL6/LIX/GCP-2, CXCL16, Cardiac Troponin I (cTnI), α2-Macroglobulin, AGP, CRP, Fetuin A, Fibrinogen, Haptoglobin, L-Selectin, PF4, SAP |
| *Cytokines and chemokines* | sCD40L, EGF, Eotaxin, FGF-2, Flt-3 ligand, Fractalkine, G-CSF, GM-CSF, GROα, IFNα2, IFNγ, IL-1α, IL-1β, IL-1ra, IL-2, IL-3, IL-4, IL-5, IL-6, IL-7, IL-8, IL-9, IL-10, IL-12p40, IL-12p70, IL-13, IL-15, IL-17A, IL-17E/IL-25, IL-17F, IL-18, IL-22, IL-27, IP-10, MCP-1, MCP-3, M-CSF, MDC (CCL22), MIG, MIP-1α, MIP-1β, PDGF-AA, PDGF-AB/BB, RANTES, TGFα, TNFα, TNFβ, VEGF-A |
| *Soluble Cytokine Receptor* | sCD30, sEGFR, sgp130, sIL-1RI, sIL-1RII, sIL-2RA, sIL-4R, sIL-6R, sRAGE, sTNFRI, sTNFRII, sVEGFR1, sVEGFR2, sVEGFR3 |

### Statistical Analysis

Demographic characteristics between the post-HDP and post-Normal pregnancy participants were compared using χ^2^ tests, Student’s T-tests, and Wilcoxon rank-sum tests. Differences in CVD-associated proteins, immune-related cytokines, and soluble cytokine receptors between the two groups were examined using Wilcoxon rank-sum tests. The log_2_(fold-change) between post-HDP and post-normal pregnancy samples were plotted against significance from Wilcoxon rank-sum tests in a Volcano plot. A Bonferroni corrected p-value of <0.0006 was considered significant for individual tests between post-HDP/post-Normal and each protein. After identifying the top four most significantly elevated proteins, we performed linear regression to examine the association between HDP status (post-HDP vs. post-Normal) and protein levels. We first completed univariable linear regression and then used multivariable linear regression adjusted for maternal age at sample collection, BMI, diabetes, and chronic hypertension. For each variable we determined the univariate (unadjusted) or multivariate coefficient, reported as the log_10_(coefficient).

Global patterns among CVD-associated proteins were investigated using Z-scores and principal components analysis. Summary Z-scores were calculated by summing Z-scores for all 15 CVD-associated proteins among all individuals and compared between post-HDP and post-Normal pregnancy participants using a two-sided Wilcoxon rank sum test. We assessed similarity of samples by constructing a heatmap and clustering samples using default settings in *pheatmap*, which utilizes an agglomerative hierarchical clustering algorithms.^39^ In short, the global expression profile of measured proteins were compared to determine how similar samples were. We then simulated data to build an empiric distribution of the number of post-HDP samples we would expect from random data to compare to the distribution in our observed samples. To quantify intragroup heterogeneity, we completed Pearson correlation by assessing expression of all proteins of interest in one sample relative to each other sample within the group of interest (e.g. compare protein expression of all CVD proteins in patient 1 to patient 2, and so forth). All statistical analyses were completed in R Studio Version 2023.09.1+494 utilizing base functions along with the packages *tidyr*,^40^ *PCAtools*,^41^ *ggplot2*,^42^ *ggfortify*,^43^ *broom*,^44^ and *wesanderson*.^45^ A p-value less than 0.05 was considered statistically significant for linear regression analyses and summary measures compared across post-normal and post-HDP groups. Similar analyses were used to examine the patterns and clustering for immune-related cytokines.

In a secondary analysis, we examined the association between protein levels of Fibrinogen and L-selectin and time interval since last pregnancy. The mean plasma protein concentration per group (post-HDP or post-Normal) was taken at each time point and the correlation with time interval since last pregnancy were examined using Pearson correlation coefficients.

## RESULTS

### Study cohort demographics

The overall study design is outlined in **Figure 1A**. We included a total of 65 participants (n=22 post-HDP and n=43 post-Normal pregnancy). There were no significant differences between groups with respect to time since pregnancy, maternal age, body mass index (BMI), rates of chronic hypertension or diabetes (**Table 2**, **Figure 1B-D)**. As expected, those with HDP delivered at earlier gestational ages and infants of lower birthweight (**Table 2**).

**Figure 1.**
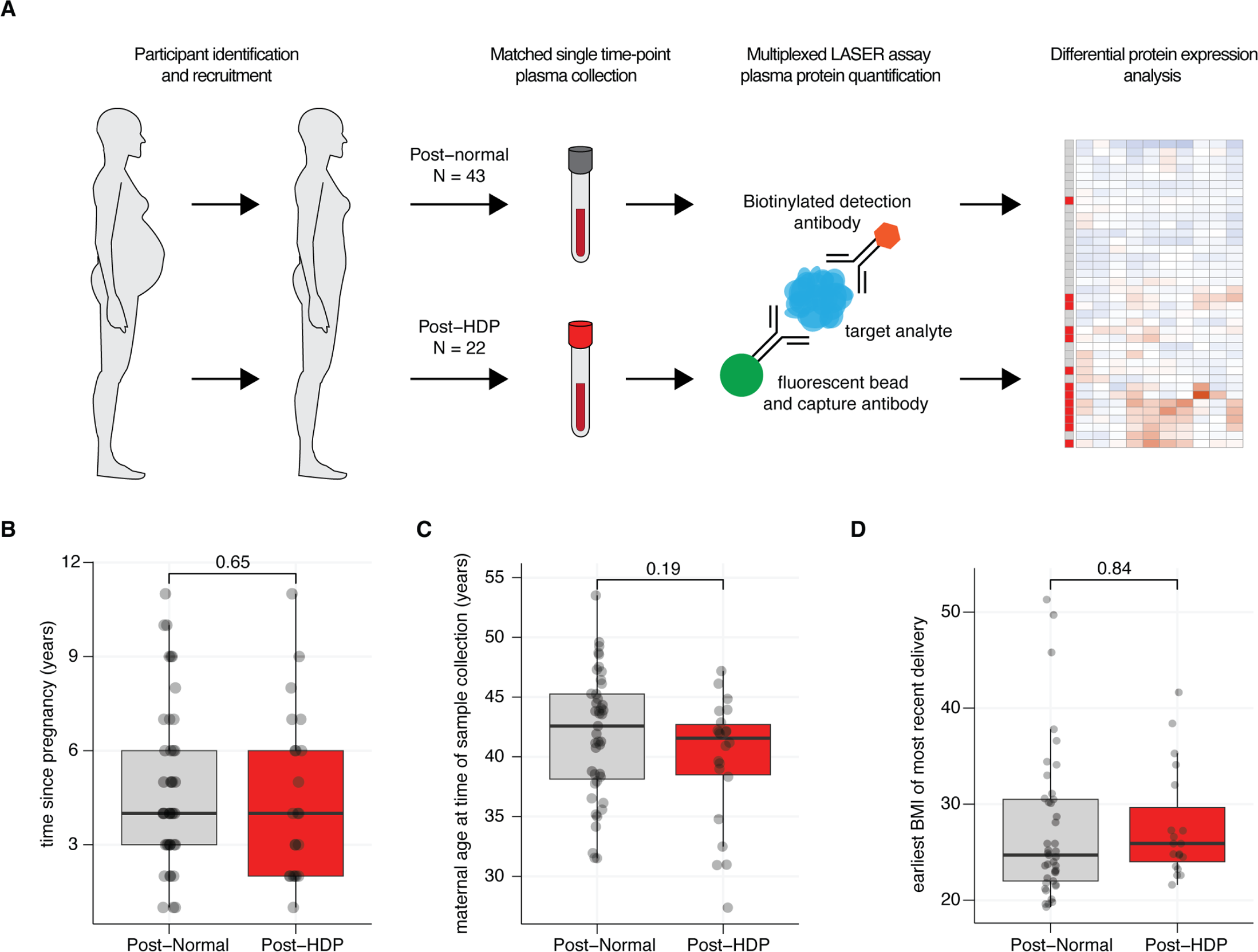
Workflow and relevant clinical parameters of the study cohort. (**A**) Schematic of sample collection, multiplex assay, and analysis. Samples were collected at a single time point and matched for time since last pregnancy. Plasma was submitted for multiplexed LASER assay protein quantification included in pre-assembled panels. (**B-D**) Boxplots comparing time since last pregnancy (**B**), maternal age at time of sample collection (**C**), and earliest body mass index (BMI) of most recent delivery (**D**) between post-Normal and post-HDP groups. For all, the *P* value is from a two-sided Wilcoxon-Rank Sum test.

**Table 2:** Demographics of the study population.

|  | Post-HDP<br>(n=22) | Post-Normal<br>(n=43) | *p*-value |
| --- | --- | --- | --- |
| Age at sample collection<br>*Missing: n=0* | 39.7 ± 5.3 | 41.8 ± 5.3 | 0.14 |
| White race<br>*Missing: n=0* | 19 (86.4) | 35 (81.4) | 0.65 |
| Hispanic ethnicity<br>*Missing: n=1* | 2 (4.6%) | 4 (18.2) | 0.16 |
| Parity<br>*Missing: n=0* | 2 (1-2) | 2 (1-2) | 0.43 |
| Time since most recent pregnancy (years)<br>*Missing: n=0* | 4 (2-6) | 4 (3-6) | 0.54 |
| Time since HDP pregnancy (years)<br>*Missing: n=0* | 6 (4-8) | N/A | N/A |
| Chronic hypertension<br>*Missing: n=0* | 3 (13.6) | 5 (11.6) | 0.82 |
| Diabetes<sup>*</sup><br>*Missing: n=0* | 4 (18.2) | 6 (13.9) | 0.44 |
| Body mass index (kg/m<sup>2</sup>)<sup>†</sup><br>*Missing: n=5* | 25.9 (23.5 – 32.0) | 24.7 (22.0 – 30.5) | 0.32 |
| Gestational age of delivery (weeks)<sup>†</sup><br>*Missing: n=2* | 37.6 (32.4 – 39.0) | 39.6 (38.6 – 40.1) | <0.001 |
| Neonatal birthweight (grams)<sup>†</sup><br>*Missing: n=1* | 2572 (1877 – 3198) | 3287 (3049 – 3651) | <0.01 |
| Cesarean delivery<sup>†</sup><br>*Missing: n=0* | 12 (54.5) | 18 (41.9) | 0.33 |
| Female neonatal sex<sup>†</sup><br>*Missing: n=0* | 10 (45.4) | 18 (41.9) | 0.78 |
| Type of HDP<br>*Missing: n=0* |  |  |  |
| Gestational hypertension | 5 (22.7) | N/A | N/A |
| Preeclampsia without severe features | 3 (13.6) | N/A | N/A |
| Preeclampsia with severe features<sup>‡</sup> | 12 (54.5) | N/A | N/A |
| Postpartum preeclampsia | 2 (9.1) | N/A | N/A |

Data are N (%), mean ( ± standard deviation), median (interquartile range)
HDP, hypertensive disorders of pregnancy
\*, includes history of gestational diabetes (in any pregnancy), type 1 diabetes, or type 2 diabetes.
†, of HDP pregnancy for the Post-HDP group and of the most recent pregnancy beyond 20 weeks
gestation for the Post-Normal group.
‡, includes superimposed preeclampsia and HELLP (hemolysis, elevated liver enzymes, low platelet)
syndrome.

### CVD associated proteins are elevated following an HDP pregnancy

Among all 77 measured proteins, fourteen proteins had significantly higher expression in the post-HDP group (**Figure 2A; Supp. Table 1**). Among these enriched proteins, the top four proteins – fibrinogen, fetuin-A36, AGP (alpha-1-acid glycoprotein), and L-selectin – are all CVD associated proteins (**Figure 2B-E**).

**Figure 2.**
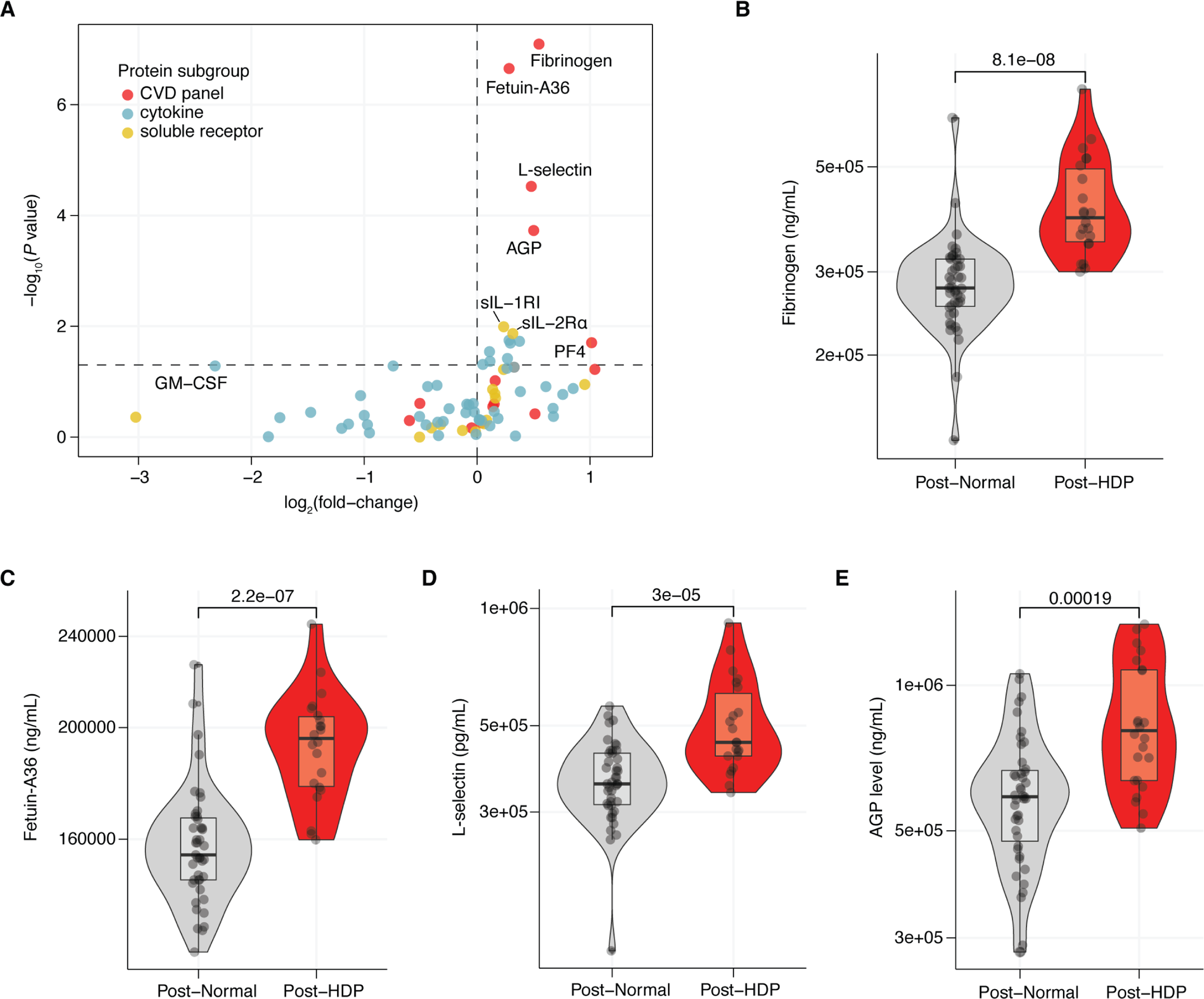
Several CVD-associated proteins are elevated in plasma in the few years after an HDP pregnancy. (**A**) Volcano plot of all 77 assays plasma proteins colored by protein subset (red = CVD panel, blue = cytokine, and yellow = soluble receptor) with the log_2_(fold-change) in protein levels between post-HDP and post-Normal and the -log_10_(*P* value) from a two-sided Wilcoxon Rank Sum Test. (**B-E**) Violin plots of fibrinogen (**B**), fetuin-A36 (**C**), L-selectin (**D**), and AGP (**E**) protein levels in plasma from individuals following a normotensive pregnancy (Post-Normal) or pregnancy complicated by HDP (Post-HDP). For all, the *P* value is from a two-sided Wilcoxon Rank Sum Test.

For the top enriched plasma proteins (fibrinogen, fetuin-A36, L-selectin, AGP), a history of HDP was significantly associated with plasma protein levels after adjustment for maternal age at sample collection, earliest BMI of the most recent pregnancy, chronic hypertension, and diabetes (**Supp. Figure 1A-D**). The estimated coefficient for post-HDP was similar between the univariable and multivariable analysis for each protein, suggesting that HDP history largely explains the elevated plasma protein levels observed in our cohort (**Supp. Figure 1E-H; Supp. Table 2**).

### Globally, CVD-associated proteins are significantly elevated in the post-HDP samples

In addition to the individual proteins listed above, we also investigated global patterns among all the CVD-associated proteins (n=15) in our cohort. Heatmap of Z-scores (**Figure 3A**) and principal component analysis (PCA) (**Figure 3B**) of the CVD-associated proteins demonstrates reasonable, but not perfect, separation of the population based on HDP history. A summary Z-score demonstrated that CVD-associated proteins are globally elevated in the post-HDP group compared to the post-Normal participants (p<0.0001) (**Figure 3C**). This indicates that exposure to HDP has variable effects on plasma protein levels; yet individuals that experienced an HDP display significant elevations in CVD-associated plasma proteins in the few years after an HDP pregnancy.

**Figure 3.**
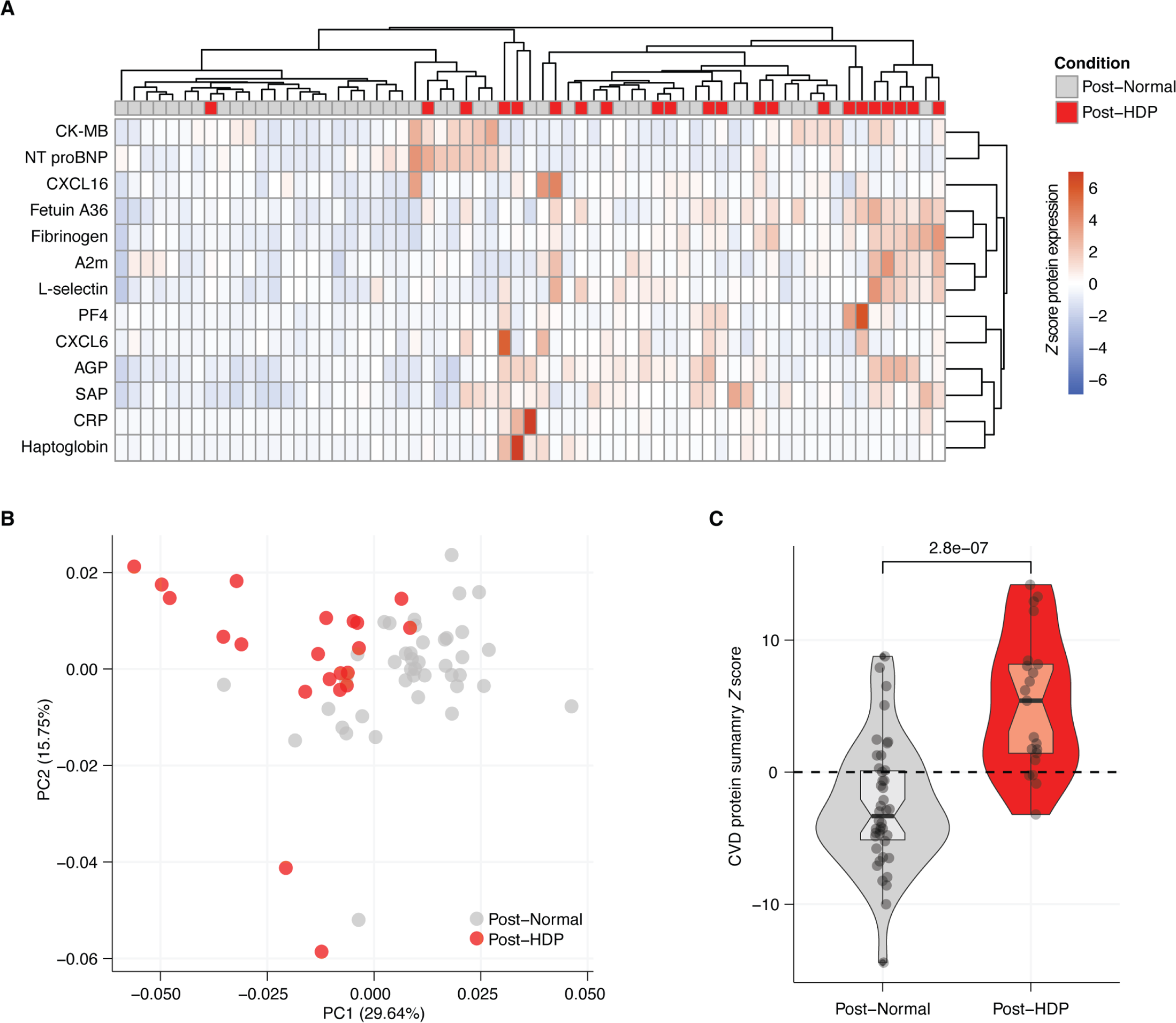
Global patterns of CVD-associated proteins in the few years after an HDP or normotensive pregnancy. (**A**) Heatmap of Z scores of CVD panel protein expression with rows reflecting an individual protein and columns corresponding to an individual participant sample. Samples colored by condition as Post-Normal (gray) or post-HDP (red). (**B**) PCA plot of Post-Normal (gray) and Post-HDP (red) protein values for plasma protein expression values of 15 CVD-associated proteins. (**C**) Violin plot of CVD summary *Z* score calculated by summing *Z* scores of all 15 CVD associated protein levels in plasma from individuals following a normotensive pregnancy (Post-Normal) or pregnancy complicated by HDP (Post-HDP). *P* value from a two-sided Wilcoxon Rank Sum Test.

### Significant heterogeneity exists among post-HDP samples

When comparing global patterns in protein expression, we observed substantial heterogeneity in CVD-associated protein levels among individual samples (**Figure 3A-B**). Simulated data that clustered the samples first split the 66 analyzed samples into two groups of 33 samples, where cluster 1 contained 15.5% (5/33) and cluster 2 contained 51.5% (17/33) post-HDP samples, respectively. We then built an empiric distribution of what fraction of post-HDP samples we would expect to see if 33 samples were selected at random (**Supp. Figure 2A-B**). The observed results were more extreme than 99.9% and 99.7% of all simulated data, for cluster 1 and cluster 2, respectively, strongly suggesting the clustering we observe is not due to random chance (**Supp. Figure 2B**).

We then quantified levels of heterogeneity amongst measured samples. We completed pairwise Pearson correlations of the plasma concentrations of CVD-associated proteins comparing each individual plasma sample to every other plasma sample in our study (**Supp. Figure 2C**). We found that post-Normal samples were significantly more similar to each other, reflected in a more positive average Pearson correlation coefficient (*R*), than post-HDP samples were to each other (**Supp. Figure 2D**).

### CVD-associated proteins in the period after pregnancy

As part of a secondary analysis, we evaluated the relationship between time since most recent pregnancy and plasma protein levels by quantifying the mean plasma protein concentration per group (post-Normal or post-HDP) at each year. We observed a significant negative correlation between fibrinogen levels and interval since last pregnancy among post-HDP participants, but not in those with a history of a normotensive pregnancy (**Supp. Figure 3A**). In comparison, plasma L-selectin levels negatively correlated with time since last pregnancy in both groups; however, levels were consistently higher in individuals with a history of HDP compared to those with a history of normotensive pregnancies (**Suppl. Figure 3B**). Other CVD-proteins did not demonstrate any notable differences across time (data not shown).

### Inflammatory markers did not vary in the few years after HDP

We did not detect circulating inflammatory or soluble cytokine receptors to be as substantially altered in the few years following HDP (**Figure 2A**). Several proteins were higher in the post-HDP population (sIL-1RI, sIL-2Rα, M-CSF, RANTES, MDC, and IL-12p40), but they did not reach significance after adjusting for multiple comparisons (**Supp. Figure 4A-F**). When assessing global patterns of inflammatory protein expression in plasma samples, we did not detect obvious clustering of post-HDP samples via dendogram or using principal component analysis (**Supp. Figure 4G-I**). Though no inflammatory proteins were significantly elevated (after adjustment for multiple comparisons), a summary *Z* score measuring the aggregate expression of all inflammatory proteins was significantly elevated in in the post-HDP samples compared to post-Normal samples (Supplementary figure 4I). This suggests that while few of the measured inflammatory proteins are significantly elevated in the few years following HDP, overall levels of inflammation may be significantly elevated in the few years following HDP.

## DISCUSSION

Here we identify several plasma proteins elevated in the few years following an HDP pregnancy. The most significantly enriched are those relevant to CVD (fibrinogen, fetuin-A, L-selectin, and AGP). In several vascular disease pathologies, high fibrinogen predicts poor outcomes, including in peripheral arterial disease, myocardial infarction, and ischemic stroke.^46^ Higher levels of fetuin-A was associated with increased risk of ischemic stroke and myocardial infarction in a relatively young population (<57 years old).^47^ Interestingly, at the same plasma level of fetuin-A, the risk of these adverse cardiovascular outcomes was higher among women than men.^47^ Although elevated L-selectin levels are associated with smoking,^48^ prior studies have noted mixed results regarding an association with CVD.^49,50^ It is, however, proposed that circulating L-selectin may serve an anti-inflammatory function with respect to atherosclerotic plaque formation.^49^ AGP, also known as orosomucoid, is associated with the presence of carotid plaques,^51^ ischemic stroke,^51,52^ and arterial stiffness.^53^ We have identified some of these same circulating markers to be elevated in the few years (median 4 years) after an HDP pregnancy.

Emerging data indicates that the HDP pregnancy itself may confer risk towards later-life CVD.^5,7,8,11^ Additionally, individuals with HDP history develop CVD along an accelerated pathway with events occurring more frequently in their 5^th^ and 6^th^ decades compared to their counterparts.^5^ Our data suggests that after accounting for several critical risk factors associated with CVD (obesity, diabetes, chronic hypertension) that those with a history of HDP demonstrate an adverse pre-clinical cardiovascular profile that may be a harbinger of increased later-life CVD risk.

We did not observe differences in individual inflammatory markers; however, persistent inflammation following an HDP pregnancy is reported in some studies.^37^ It is possible that we were underpowered to detect differences and that these results require further contextualization with cellular immune data. When assessing inflammatory markers in aggregate, we did detect a significant elevation in post-HDP samples compared to controls. Collectively, this could indicate persistent low-level inflammation following HDP, or elevations in inflammatory markers not included in our study. Our results, however, suggest that CVD-associated proteins may be more robust biomarkers than inflammatory mediators for inclusion in future prospective studies.

Few studies have investigated circulating biomarkers in the years after the HDP pregnancy. In those with HDP, one study demonstrated increased TNF-ɑ three months postpartum^54^ and another noted an increased IL-6/IL-10 ratio 20 years after the index pregnancy.^55^ A recent study that evaluated some cardiovascular (VCAM-1, VEGF, CD40L, GDF-15, and ST-2) and metabolic biomarkers in individuals 5-10 years after index pregnancy did not find an association between HDP and these biomarkers.^56^ We also noted no difference in some of these same markers (VCAM-1, VEGF, CD40L), however, uncovered well-established CVD-associated proteins (fibrinogen, fetuin-A, L-selectin, AGP) to be elevated in the few years following HDP that were not measured in this above-listed study. We also observed heterogeneity in the levels of these CVD-associated proteins in the post-HDP cohort. This may support a more recent understanding of multiple phenotypes of preeclampsia giving insight into the possibility of developing a unique CVD outcome based on the HDP phenotype.^57^

Our study is not without limitations. We only had access to single time point samples, thus our analysis of the changes in plasma protein levels by interval from pregnancy (**Suppl. Figure 3A-B**) requires validation with serial sampling from the same individual. For example, fibrinogen is an acute phase reactant,^58–61^ and as our participants presented as outpatients, non-pregnant, and without evidence of any acute illness, further work is needed to clarify the etiology of this elevation in those with a history of HDP in the absence of known causes. We also lack pre-pregnancy measures to more definitively determine whether the HDP exposure plays a role in leading to increased levels of CVD-associated proteins. Conversely, pre-pregnancy measures are challenging to obtain as recruitment of thousands of individuals are necessary to capture enough HDP cases, particularly those with severe phenotypes carrying the highest risk for later-life CVD. Lastly, our population lacks adequate representation from American Indian and Alaskan Native and African American pregnant individuals, acknowledging that they experience the highest rates of HDP in the United States (US).^62^

Strengths of our study include use of a highly sensitive multiplex assay that can measure numerous proteins with minimal sample volume. Employment of this expansive approach in this rare cohort type is necessary as foundational data regarding post-HDP biology is drastically lacking. We were not subject to batch effects as all samples were run simultaneously. We employed robust statistical methods, including corrected p-values for multiple comparisons, and orthogonal accounting for underlying cardiovascular risk by controlling for age, BMI, diabetes, and chronic hypertension. Although we found significant differences in several key CVD-associated proteins, additional studies in larger cohorts are needed to further advance our understanding of post-HDP cardiovascular biology.

### Perspectives

The prevalence of hypertensive disorders of pregnancy, such as preeclampsia, is on the rise. The prevalence of HDP among US delivery hospitalizations in 2019 was close to 16%, up from 13% just two years prior.^62^ At the other end of the life course, CVD remains the leading worldwide cause of mortality in women.^1^ With the well-established connection between HDP and CVD and the combined global burdens of these two entities, stratification of cardiovascular risk in the intervening years, after HDP but before evident CVD, is urgently needed. We present the first cohort demonstrating significant elevations, by orders of magnitude, in several key proteins known to be implicated in CVD in the few years following an HDP pregnancy compared to matched participants, and after controlling for critical confounders. This provides molecular evidence of post-HDP cardiovascular risk and that exposure to the HDP pregnancy itself may have measurable impact. These circulating biomarkers may represent promising targets for identifying those at highest risk for developing CVD and informing interventions that mitigate CVD event risk in this high-risk population.

### Novelty and Relevance

#### 1. What Is New?

- Several circulating proteins implicated in cardiovascular disease (CVD) are considerably elevated in the few years after a hypertensive disorder of pregnancy (HDP).
- Although HDP is characterized by widespread inflammation, individual inflammatory markers are only modestly elevated in the few years after HDP.
- We provide molecular evidence of post-HDP cardiovascular risk and that the exposure to the HDP pregnancy itself may have measurable impact.

#### 2. What Is Relevant?

- The prevalence of HDP is on the rise and HDP is strongly linked to an increased risk for later-life CVD.
- The combined global burdens of HDP and CVD make understanding mechanisms contributing to cardiovascular risk in the few years after HDP critical.

#### 3. Clinical/Pathophysiological Implications?

- This study raises questions about whether the HDP pregnancy itself confers risk, in line with epidemiologic data which supports this notion and indicates that individuals with HDP history develop CVD along an accelerated pathway.

## ACKNOWLEDGEMENTS

None

## SOURCES OF FUNDING

NIH/NHLBI K08HL150169, NIH/NICHD R21HD086620 (R. Shree)

NIH/NICHD K12HD000849, Burroughs Wellcome Fund Grant# 1288787, Doris Duke Clinical Scientist Development Award #2023-0231 (S. McCartney)

NIH/NCATS UL1 TR002319 (University of Washington)

These funders had no role in the design or conduct of this study, nor in the interpretation of results.

## REFERENCES

1. Virani Salim S., Alonso Alvaro, Aparicio Hugo J., et al. Heart Disease and Stroke Statistics—2021 Update. Circulation. 2021;143(8):e254–e743.

2. Lawton JS. Sex and gender differences in coronary artery disease. Semin Thorac Cardiovasc Surg. 2011;23(2):126–130.

3. Bairey Merz CN, Shaw LJ, Reis SE, et al. Insights from the NHLBI-Sponsored Women’s Ischemia Syndrome Evaluation (WISE) Study: Part II: gender differences in presentation, diagnosis, and outcome with regard to gender-based pathophysiology of atherosclerosis and macrovascular and microvascular coronary disease. J Am Coll Cardiol. 2006;47(3 Suppl):S21–29.

4. Melchiorre K, Thilaganathan B, Giorgione V, Ridder A, Memmo A, Khalil A. Hypertensive Disorders of Pregnancy and Future Cardiovascular Health. Front Cardiovasc Med. 2020;7:59.

5. Honigberg MC, Zekavat SM, Aragam K, et al. Long-Term Cardiovascular Risk in Women With Hypertension During Pregnancy. J Am Coll Cardiol. 2019;74(22):2743–2754.

6. Honigberg MC, Zekavat SM, Raghu VK, Natarajan P. Microvascular Outcomes in Women With a History of Hypertension in Pregnancy. Circulation. 2022;145(7):552–554.

7. de Havenon A, Delic A, Stulberg E, et al. Association of Preeclampsia With Incident Stroke in Later Life Among Women in the Framingham Heart Study. JAMA Netw Open. 2021;4(4):e215077.

8. Hansen AL, Søndergaard MM, Hlatky MA, et al. Adverse Pregnancy Outcomes and Incident Heart Failure in the Women’s Health Initiative. JAMA Netw Open. 2021;4(12):e2138071.

9. Brown MC, Best KE, Pearce MS, Waugh J, Robson SC, Bell R. Cardiovascular disease risk in women with pre-eclampsia: systematic review and meta-analysis. Eur J Epidemiol. 2013;28(1):1–19.

10. Rich-Edwards JW, McElrath TF, Karumanchi SA, Seely EW. Breathing life into the lifecourse approach: pregnancy history and cardiovascular disease in women. Hypertens Dallas Tex 1979. 2010;56(3):331–334.

11. Romundstad PR, Magnussen EB, Smith GD, Vatten LJ. Hypertension in pregnancy and later cardiovascular risk: common antecedents? Circulation. 2010;122(6):579–584.

12. Mosca L, Benjamin EJ, Berra K, et al. Effectiveness-based guidelines for the prevention of cardiovascular disease in women--2011 update: a guideline from the American Heart Association. Circulation. 2011;123(11):1243–1262.

13. Lewey J, Beckie TM, Brown HL, et al. Opportunities in the Postpartum Period to Reduce Cardiovascular Disease Risk After Adverse Pregnancy Outcomes: A Scientific Statement From the American Heart Association. Circulation. 2024;149(7):e330–e346.

14. Noori M, Donald AE, Angelakopoulou A, Hingorani AD, Williams DJ. Prospective study of placental angiogenic factors and maternal vascular function before and after preeclampsia and gestational hypertension. Circulation. 2010;122(5):478–487.

15. Mori T, Watanabe K, Iwasaki A, et al. Differences in vascular reactivity between pregnant women with chronic hypertension and preeclampsia. Hypertens Res Off J Jpn Soc Hypertens. 2014;37(2):145–150.

16. Henriques ACPT, Carvalho FHC, Feitosa HN, Macena RHM, Mota RMS, Alencar JCG. Endothelial dysfunction after pregnancy-induced hypertension. Int J Gynaecol Obstet Off Organ Int Fed Gynaecol Obstet. 2014;124(3):230–234.

17. Hamad RR, Eriksson MJ, Silveira A, Hamsten A, Bremme K. Decreased flow-mediated dilation is present 1 year after a pre-eclamptic pregnancy. J Hypertens. 2007;25(11):2301–2307.

18. Barnes JN, Harvey RE, Miller KB, et al. Cerebrovascular Reactivity and Vascular Activation in Postmenopausal Women With Histories of Preeclampsia. Hypertens Dallas Tex 1979. 2018;71(1):110–117.

19. Ehrenthal DB, Goldstein ND, Wu P, Rogers S, Townsend RR, Edwards DG. Arterial stiffness and wave reflection 1 year after a pregnancy complicated by hypertension. J Clin Hypertens Greenwich Conn. 2014;16(10):695–699.

20. Yuan LJ, Xue D, Duan YY, Cao TS, Yang HG, Zhou N. Carotid arterial intima–media thickness and arterial stiffness in pre-eclampsia: analysis with a radiofrequency ultrasound technique. Ultrasound Obstet Gynecol Off J Int Soc Ultrasound Obstet Gynecol. 2013;42(6):644–652.

21. Lampinen KH, Rönnback M, Kaaja RJ, Groop PH. Impaired vascular dilatation in women with a history of pre-eclampsia. J Hypertens. 2006;24(4):751–756.

22. Blaauw J, Souwer ETD, Coffeng SM, et al. Follow up of intima-media thickness after severe early-onset preeclampsia. Acta Obstet Gynecol Scand. 2014;93(12):1309–1316.

23. Akhter T, Larsson M, Wikström AK, Naessen T. Thicknesses of individual layers of artery wall indicate increased cardiovascular risk in severe pre-eclampsia. Ultrasound Obstet Gynecol Off J Int Soc Ultrasound Obstet Gynecol. 2014;43(6):675–680.

24. Aykas F, Solak Y, Erden A, et al. Persistence of cardiovascular risk factors in women with previous preeclampsia: a long-term follow-up study. J Investig Med Off Publ Am Fed Clin Res. 2015;63(4):641–645.

25. Shahul S, Rhee J, Hacker MR, et al. Subclinical left ventricular dysfunction in preeclamptic women with preserved left ventricular ejection fraction: a 2D speckle-tracking imaging study. Circ Cardiovasc Imaging. 2012;5(6):734–739.

26. Melchiorre K, Sutherland GR, Liberati M, Thilaganathan B. Preeclampsia is associated with persistent postpartum cardiovascular impairment. Hypertens Dallas Tex 1979. 2011;58(4):709–715.

27. Haas David M., Parker Corette B., Marsh Derek J., et al. Association of Adverse Pregnancy Outcomes With Hypertension 2 to 7 Years Postpartum. J Am Heart Assoc. 2019;8(19):e013092.

28. Field C, Grobman WA, Wu J, et al. Elevated Blood Pressure in Pregnancy and Long-Term Cardiometabolic Health Outcomes. Obstet Gynecol. Published online May 5, 2022:10.1097/AOG.0000000000005674.

29. Stanhewicz AE, Jandu S, Santhanam L, Alexander LM. Alterations in endothelin type B receptor contribute to microvascular dysfunction in women who have had preeclampsia. Clin Sci Lond Engl 1979. 2017;131(23):2777–2789.

30. Stanhewicz AE, Jandu S, Santhanam L, Alexander LM. Increased Angiotensin II Sensitivity Contributes to Microvascular Dysfunction in Women Who Have Had Preeclampsia. Hypertens Dallas Tex 1979. 2017;70(2):382–389.

31. Saito S, Sakai M. Th1/Th2 balance in preeclampsia. J Reprod Immunol. 2003;59(2):161–173.

32. Rana S, Lemoine E, Granger JP, Karumanchi SA. Preeclampsia. Circ Res. 2019;124(7):1094–1112.

33. Alfaddagh A, Martin SS, Leucker TM, et al. Inflammation and cardiovascular disease: From mechanisms to therapeutics. Am J Prev Cardiol. 2020;4:100130.

34. Bassuk SS, Rifai N, Ridker PM. High-sensitivity C-reactive protein: clinical importance. Curr Probl Cardiol. 2004;29(8):439–493.

35. Miller VM, Redfield MM, McConnell JP. Use of BNP and CRP as biomarkers in assessing cardiovascular disease: diagnosis versus risk. Curr Vasc Pharmacol. 2007;5(1):15–25.

36. Diederichsen MZ, Diederichsen SZ, Mickley H, et al. Prognostic value of suPAR and hs-CRP on cardiovascular disease. Atherosclerosis. 2018;271:245–251.

37. Herrock O, Deer E, LaMarca B. Setting a stage: Inflammation during preeclampsia and postpartum. Front Physiol. 2023;14:1130116.

38. Bowser BL, Robinson RAS. Enhanced Multiplexing Technology for Proteomics. Annu Rev Anal Chem. 2023;16(Volume 16, 2023):379–400.

39. Kolde R. pheatmap: Pretty Heatmaps. Published online January 4, 2019.

40. Wickham H, Vaughan D, Girlich M, Ushey K, Software P, PBC. tidyr: Tidy Messy Data. Published online January 24, 2024.

41. Blighe K. kevinblighe/PCAtools. Published online July 17, 2024.

42. Wickham H. Ggplot2. Springer International Publishing; 2016.

43. Tang Y, Horikoshi M, Li W. ggfortify: Unified Interface to Visualize Statistical Results of Popular R Packages. R J. 2016;8(2):474–485.

44. Robinson D, Hayes A, Couch [aut S, et al. broom: Convert Statistical Objects into Tidy Tibbles. Published online May 17, 2024.

45. Ram K, Wickham H, Richards C, Baggett A. wesanderson: A Wes Anderson Palette Generator. Published online October 31, 2023.

46. Surma S, Banach M. Fibrinogen and Atherosclerotic Cardiovascular Diseases—Review of the Literature and Clinical Studies. Int J Mol Sci. 2022;23(1):193.

47. Weikert C, Stefan N, Schulze MB, et al. Plasma fetuin-a levels and the risk of myocardial infarction and ischemic stroke. Circulation. 2008;118(24):2555–2562.

48. Scott D, Palmer R. The influence of tobacco smoking on adhesion molecule profiles. Tob Induc Dis. 2002;1(1):3.

49. Berardi C, Decker PA, Kirsch PS, et al. Plasma and serum L-selectin and clinical and subclinical cardiovascular disease: the Multi-Ethnic Study of Atherosclerosis (MESA). Transl Res J Lab Clin Med. 2014;163(6):585–592.

50. Wei YS, Lan Y, Meng LQ, Nong LG. The association of L-selectin polymorphisms with L-selectin serum levels and risk of ischemic stroke. J Thromb Thrombolysis. 2011;32(1):110–115.

51. Berntsson J, Östling G, Persson M, Smith JG, Hedblad B, Engström G. Orosomucoid, Carotid Plaque, and Incidence of Stroke. Stroke. 2016;47(7):1858–1863.

52. Gannon BM, Glesby MJ, Finkelstein JL, Raj T, Erickson D, Mehta S. A point-of-care assay for alpha-1-acid glycoprotein as a diagnostic tool for rapid, mobile-based determination of inflammation. Curr Res Biotechnol. 2019;1:41–48.

53. Nilsson Wadström B, Persson M, Engström G, Nilsson PM. Aortic Stiffness, Inflammation, and Incidence of Cardiovascular Events in Elderly Participants From the General Population. Angiology. 2022;73(1):51–59.

54. Herrock OT, Deer E, Amaral LM, et al. B2 cells contribute to hypertension and natural killer cell activation possibly via AT1-AA in response to placental ischemia. Am J Physiol Renal Physiol. 2023;324(2):F179–F192.

55. Freeman DJ, McManus F, Brown EA, et al. Short- and Long-Term Changes in Plasma Inflammatory Markers Associated With Preeclampsia. Hypertension. 2004;44(5):708–714.

56. Battarbee AN, Mele L, Landon MB, et al. Hypertensive Disorders of Pregnancy and Long-Term Maternal Cardiovascular and Metabolic Biomarkers. Am J Perinatol. 2024;41(S 01):e1976–e1981.

57. Roberts JM, Rich-Edwards JW, McElrath TF, Garmire L, Myatt L, for the Global Pregnancy Collaboration. Subtypes of Preeclampsia: Recognition and Determining Clinical Usefulness. Hypertension. 2021;77(5):1430–1441.

58. Schupp T, Weidner K, Rusnak J, et al. Fibrinogen reflects severity and predicts outcomes in patients with sepsis and septic shock. Blood Coagul Fibrinolysis Int J Haemost Thromb. 2023;34(3):161–170.

59. De Sutter J, De Buyzere M, Gheeraert P, et al. Fibrinogen and C-reactive protein on admission as markers of final infarct size after primary angioplasty for acute myocardial infarction. Atherosclerosis. 2001;157(1):189–196.

60. Hellgren M. Hemostasis during normal pregnancy and puerperium. Semin Thromb Hemost. 2003;29(2):125–130.

61. Hamar P. Local Production of Acute Phase Proteins: A Defense Reaction of Cancer Cells to Injury with Focus on Fibrinogen. Int J Mol Sci. 2024;25(6):3435.

62. Ford ND. Hypertensive Disorders in Pregnancy and Mortality at Delivery Hospitalization — United States, 2017–2019. MMWR Morb Mortal Wkly Rep. 2022;71.

